# The herbal combination of Sugarcane, Black Myrobalan, and mastic as a supplementary treatment for COVID-19: a randomized clinical trial

**DOI:** 10.1101/2021.04.27.21256221

**Authors:** Alireza Hashemi Shiri, Esmaeil Raiatdoost, Hamid Afkhami, Ruhollah Ravanshad, Seyed Ehsan Hosseini, Navid Kalani, Rahim Raoufi

## Abstract

**Background:** Given the COVID-19 pandemic’s, researchers are beseeched for effective treatments. Herbal medicine is also queried for potential supplementary treatments for COVID-19. We aimed to evaluate the effects of Sugarcane, Black Myrobalan, and Mastic herbal medications for COVID-19 patients.

**Methods:** This was a double-blinded randomized clinical trial study conducted over three months from May to July 2020 in patients admitted with a diagnosis of COVID-19 in Peymaniyeh Hospital in Jahrom, Iran. The intervention group received the treatment protocol approved by the Ministry of Health of Iran during the period of hospitalization and the herbal supplement obtained from the combination of black myrobalan and mastic and sugarcane, twice a day (3g of herbal supplements). All patients were compared in terms of demographic variables, vital signs, clinical and laboratory variables.

**Results:** 72 patients with COVID-19, divided into intervention (n=37) and control (n=35) groups. intervention and control groups had not any significant difference in terms of baseline characteristics. The time-to-event analysis revealed a significant difference in 4 symptoms of cough, fever, dyspnea, and myalgia (P<0.05). The Control group had a significantly lower decrease in C-reactive protein during 7 days (P<0.05). Patients in the herbal supplement group were hospitalized for 4.12 days and in the control group were hospitalized for 8.37 days (P=0.001). ICU admission and death only happened in 3 (8.6%) patients of the control group.

**Conclusion:** While advanced studies with more sample size are needed; the proposed combination seems to be effective in the symptom treatment and reducing the length of hospitalization.

## Background

COVID-19 is a viral disease that has been responsible for the deaths of large numbers of people around the world in 2020. COVID-19 causes pneumonia, with classic symptoms of fever, cough, dyspnea, and myalgia (Park et al., 2020). It can have so wide range of symptoms and also damage other organs such as the heart, liver, kidneys. Some patients eventually die from multiple organ failure, shock, acute respiratory distress syndrome, heart failure, arrhythmia, and renal failure (Cevik et al., 2020). During this pandemic, various clinical trials have been launched to examine different medications, mainly with properties in strengthening the body’s immune system, antiviral, and anti-inflammatory properties to prevent cytokine storm (Maguire and Guérin, 2020). Meanwhile, various studies have shown that traditional herbal medicine could improve the symptoms of COVID-19 (Li et al., 2020). In this regard, Traditional Iranian Medicine (TIM) has potential propositions that could be taken to account as medications to improve COVID-19 symptoms. Some clinical trials are being conducted in Iran to assess the effect of herbal medications for COVID-19. One of our potential herbal candidates for this aim was the Black myrobalan (Terminalia chebula or black myrobalan) (Singh and Kumar, 2013), due to its wide range of biologically active compounds and its applications in TIM for the treatment of respiratory tract diseases (Saleem et al., 2002; Belapurkar et al., 2014). Our next candidate, sugarcane (Saccharum officinarum) has been used extensively in TIM (Singh et aa;., 2015). Its beneficial effects are supported by in vivo/vitro studies, including antihypertensive, anti-inflammatory, anti-hypertensive, and anti-hepatotoxic activity (Arruzazabala et al., 1994; Ledon et al., 2003; Jin et al., 1981). Pistacia lentiscus resina (mastics) was another herb that attracted researchers due to reducing the symptoms of autoimmune diseases by inhibiting the hyperinflammatory pathways (Dimas et al., 2012). In this study, we examined the combining of these three plants (Sugarcane, black myrobalan, and mastic) along with the treatment protocol of the Ministry of Health on COVID_19 patients.

## Methods

### Study design

The present study is a double-blind, randomized clinical trial that was conducted over three months from April 2020 to June 2020 in patients admitted with a diagnosis of COVID-19 in Peymaniyeh Hospital in Jahrom, Iran.

### Ethical considerations

Before entering the patients in this study, the research process was explained and informed consent was obtained from them. Throughout the study, researchers adhered to the principles of the Helsinki Declaration and the confidentiality of patient information. All costs of the project were covered by the researchers and no additional costs were incurred by the patients. This study was approved by the ethics committee of Jahrom University of Medical Sciences under the ethical code IR.JUMS.REC.1399.003 and was registered in the Iranian registry of clinical trials under the number IRCT20200415047082N1.

### Sampling

The study population was patients admitted with a definitive diagnosis of COVID-19 in the wards of Peymanieh Hospital in Jahrom. Sample size assuming standard difference=0.85 and confidence limits of 95% and power = 80% and assuming an equal number of samples in each group using Altman nomogram and taking into account 15% precipitation, 70 Person was determined. Then, to have an equal chance of being in the intervention group or control group, the samples were randomly assigned to the study groups using a random number table.

Inclusion criteria: Patients admitted with COVID-19 with a definitive diagnosis of PCR test, having age over 18 years, and not being pregnant or lactating. Patients with definitions of severe COVID-19, as well as severe respiratory distress syndrome, organ failure, and ICU admitted patients were not included in the study. The infectious disease specialist supervised these criteria.

Exclusion criteria: dissatisfaction with participation in the study, dissatisfaction with continuing herbal supplementation, history of severe cardiovascular disease, severe shortness of breath, uncontrolled diabetes, severe kidney or liver disease or any uncontrolled systemic disease, History of drug abuse, and current anti-psychosis (Flow diagram 1, has showed the flow chart of study sampling).

### Intervention

All patients with inclusion criteria at the time of the study, after obtaining written consent and explaining the study conditions, entered the study. patients participating in the present study were divided into intervention and control groups by tossing coins. Patients were adjusted for age and sex. The treatment protocol in the two groups of intervention and control was as follows. the intervention group received the treatment protocol approved by the Ministry of Health of Iran during the period of hospitalization and the herbal supplement obtained from the combination of black myrobalan and mastic and sugarcane, twice a day. the control group only received the approved treatment protocol. Based on a literature review, optimal doses with the lowest risk of adverse events were chosen. In the case of sugarcane, according to the American Heart Association (AHA), the permissible daily intake of sugar for women is 6 teaspoons equivalent to 25 grams, and for men, 9 teaspoons equivalent to 36 grams; in our study, 3 grams sugarcane per day (1.5 grams BID) was used based on the TIM principles, which was safe based on the AHA principles, too. Black myrobalan extract was used in a dose of the 1 gram single dose per day.

In the case of the mastic, a dose of 1 gram twice daily has been approved for the treatment of benign gastric ulcers. Also, we used 1 gram mastic twice daily in our study.

### Herbal supplement production method

To prepare the desired herbal supplement, sugarcane, mastic, and black myrobalan were purchased from approved herb suppliers. The originality of the plants was confirmed by a botanist. Plants were washed and dried to be powder by shredder considering the sterility. Powders were kept in special packages containing 3000 mg of herbal supplements (0.5 gram black myrobalans, 1 gram mastic, 1.5 grams sugarcane), which were given to patients evening and night before sleep. Based on the TIM guidance, the medication should be used sublingually and the patient had not to try swallow or chew it first but had to allow saliva to be secreted and mixed with it and gradually swallow it. Failure to pay attention to this issue causes nausea in the patient based on TIM. It should be noted that drinking water with this supplement and even up to an hour after taking it was forbidden due to the reduced effectiveness of the drug with water, so we ask the patient to help us in this matter.

### Control group

The Control group was planned to receive a placebo and the treatment protocol approved by the Ministry of Health of Iran during the period of hospitalization. Placebo was shape, size, and color-matched with the main supplement in the intervention group. It was made of bran and barley powder (for color matching).

### Blinding

Based on the randomization outcome declared by the lead nurse, the administration of herbal supplements in the intervention group, and placebo in the control group were performed by nurses. The researchers and nurses did not realize the randomization results and the type of package provided to the patient. The researcher only provided the supplement and placebo to the head nurse, in the same form of packaging.

### Data collection

All patients were compared in terms of demographic variables, vital signs, clinical and laboratory variables. Demographic characteristics included: age, gender, Body mass index (BMI), history of smoking, occupation. Critical indicators including temperature, systolic and diastolic blood pressure, heart rate, arterial blood oxygen saturation, and respiration rate were examined and recorded daily in both groups. The averaged vital sign values and daily values were compared. Vital sign of each day was recorded three times a day, including temperature (C), blood pressure (mmHg), pulse rate (beats per minute), respiratory rate (beats per minute), blood oxygen saturation (percent). Clinical characteristics including symptoms (Cough, Dyspnea, Myalgia, Fatigue, sputum discharge, Rhinorrhea, and Headache) were assessed daily with a designed questionnaire. In this questionnaire, the patient first determines the presence of these symptoms at the beginning of the disease and finally choose one of the options (It got much better, It got better, It didn’t change, It got worse, and It got much worse); symptoms tracks were recorded for each of these symptoms. Laboratory indices were performed in the first to seventh days of hospitalization. Laboratory indicators included: aspartate aminotransferase (AST), alanine aminotransferase (ALT), prothrombin time (PT), partial thromboplastin time (PTT), international normalized ratio (INR), Blood urea nitrogen (BUN), creatinine (Cr), white blood cells (WBCs), red blood cells (RBCs), hemoglobin (HB), neutrophil, lymphocyte, monocyte counts.

### Data analysis

Data analysis was performed by descriptive statistics indicators (frequency, percentage, mean and standard deviation) and inferential statistical tests (Chi-square, ANOVA, and Repeated measurement) using SPSS software version 21. The significance level was considered P <0.05.

## Results

In this study, 72 patients with COVID-19, divided into intervention (n = 37) and control (n = 35) groups were studied. There were 17 (48.6%) male subjects in the control and 19 (51.4%) male subjects in the intervention group. The results of statistical analysis showed that the intervention and control groups had not any significant difference in terms of age, sex, BMI, smoking history, and occupation (P>0.05), (Table 1).

**Table 1.**
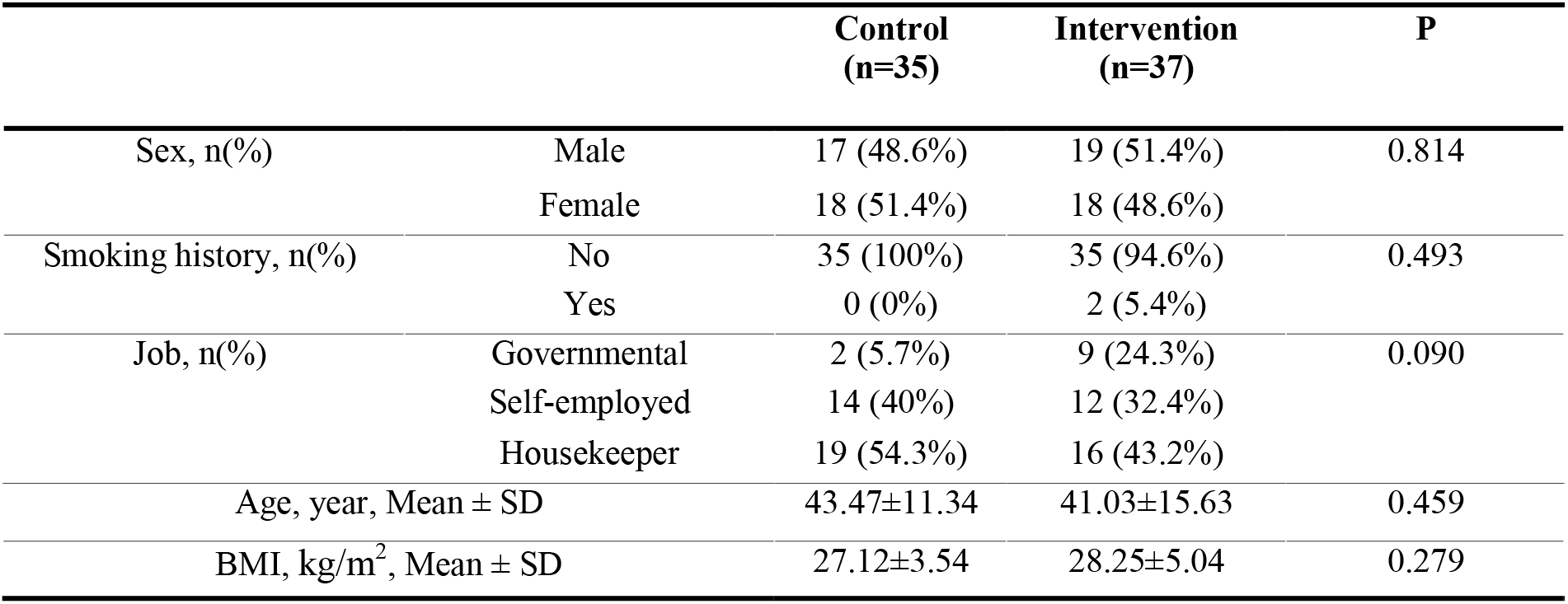
Baseline characteristics of study participants

Time to symptom disappearance was assessed for the major symptoms. The time-to-event analysis revealed a statistically significant difference in 4 symptoms of cough, fever, dyspnea, and myalgia (figure 1). where median rate ratio of the cough disappearance in intervention group versus control group was 0.285 (CI95%:0.173-0.427; P<0.05); rate ratio of the fever disappearance was 0.5 (CI95%:0.271 - 0.921; P<0.05); rate ratio of the dyspnea disappearance was 0.285 (CI95%:0.169 - 0.480; P<0.05); and it was 0.333 (CI95%:0.185 - 0.598; P<0.05) for myalgia disappearance (table 2). Detailed analysis of daily change in all symptoms is reported in supplementary table S1.

**Table 2.** Time-to-event analysis of major symptoms

|  | Control | Intervention | Rate Ratio | P |
| --- | --- | --- | --- | --- |
| Cough, Median (95% CI) | 7 (5.93 - 7.92) | 2 (1.9 - 2.57) | 0.285 (0.173- 0.427) | <0.0001 |
| Fever, Median (95% CI) | 2 (1.307 - 2.825) | 1 (0.519 - 1.222) | 0.5 (0.271 - 0.921) | 0.0051 |
| Dyspnea, Median (95% CI) | 7 (6.404 - 8.595) | 2 (1.481 - 2.518) | 0.285 (0.169 - 0.480) | <0.0001 |
| Myalgia, Median (95% CI) | 3 (1.669 - 3.463) | 1 (0.629 - 1.177) | 0.333 (0.185 - 0.598) | <0.0001 |

**Figure 1.**
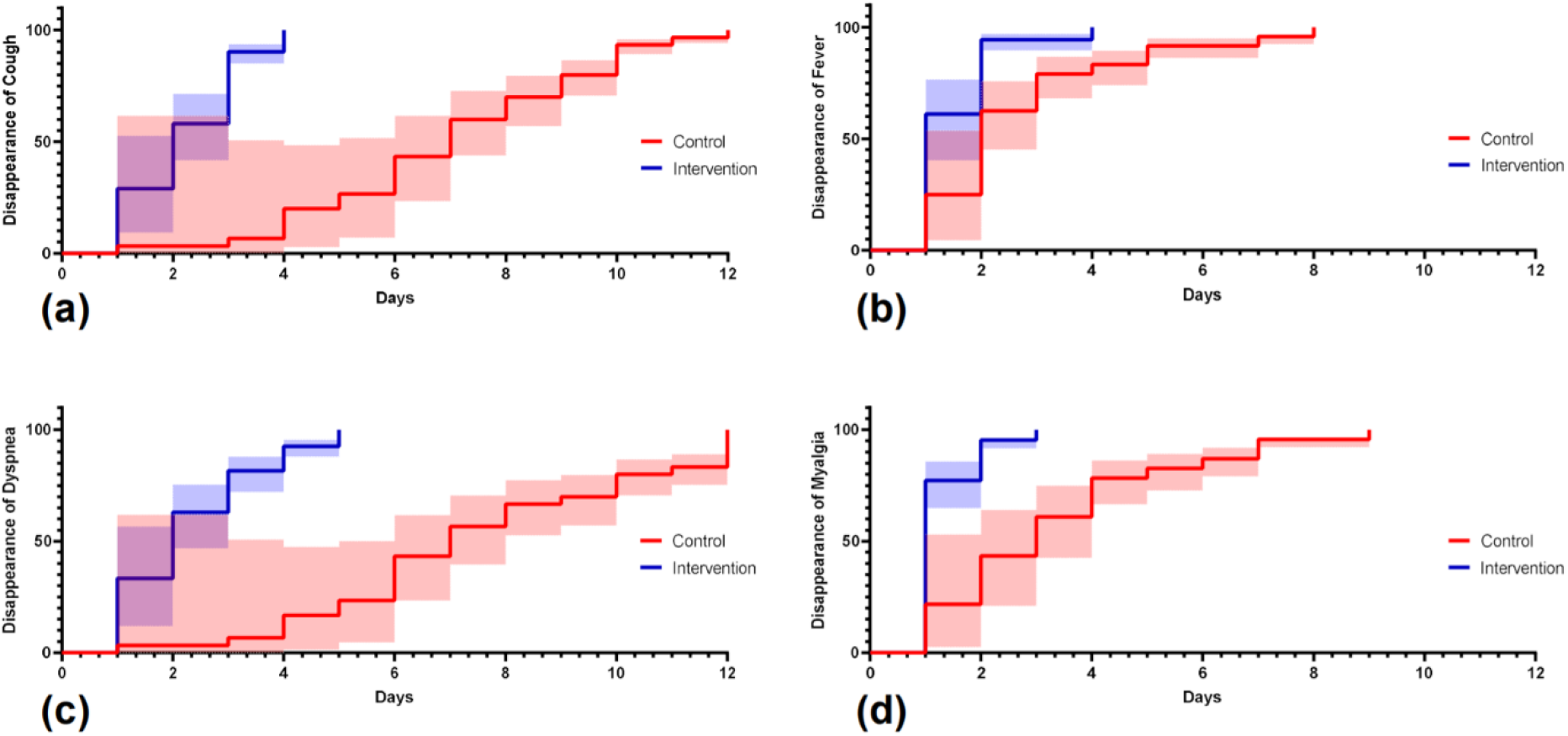
Cumulative recovery estimates of (a) cough, (b) fever, (c) dyspnea, and (d) myalgia in the control and intervention group.

**Figure 2.**
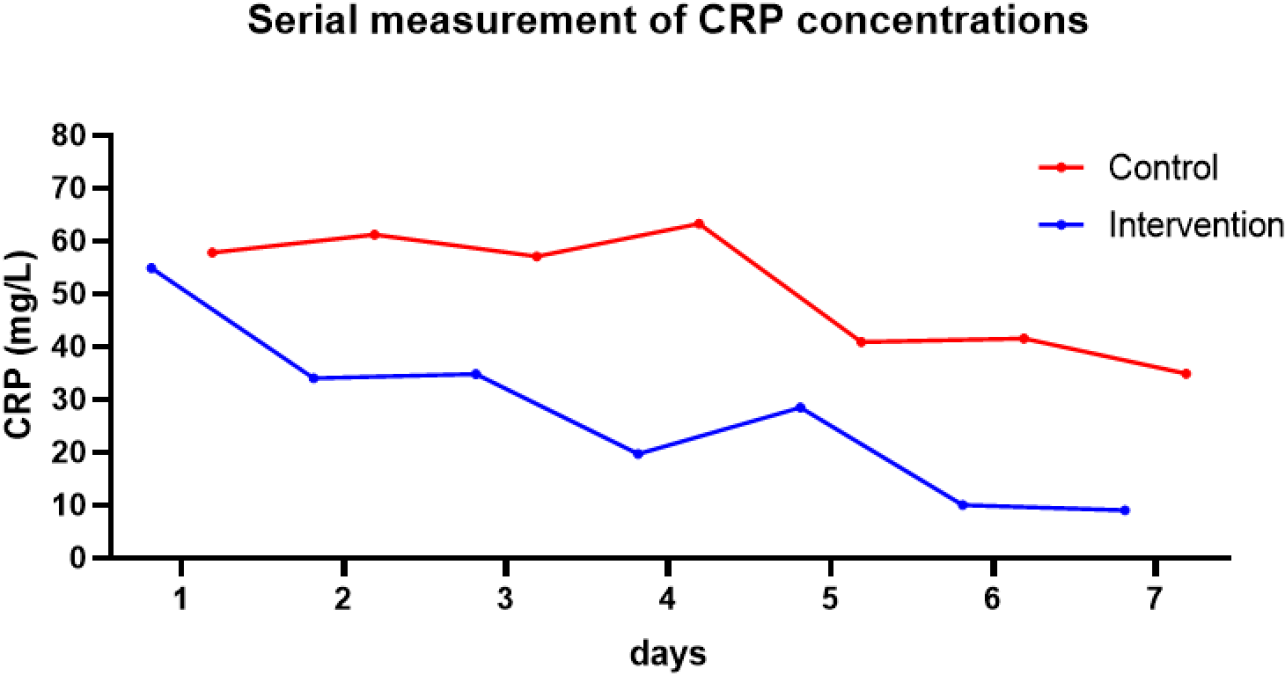
Serial measurement of CRP.

An evaluation of patients’ condition, daily vital signs were recorded and averaged value of vital signs were compared between two groups. There wasn’t any significant difference in terms of averaged O2 Saturation, systolic blood pressure (SBP), diastolic blood pressure (DBP), pulse rate (PR), and respiratory rate (RR) between the two groups (P>0.05), as shown in table 3. Daily vital signs are shown in table S2.

**Table 3.** Vital signs of study groups

|  | Control |  | Intervention |  | p-value |
| --- | --- | --- | --- | --- | --- |
|  | mean | SD | mean | SD |  |
| **O2 Saturation** | 88.9 | 0.374 | 92.061 | 1.938 | 0.1218 |
| **SBP** | 112.371 | 3.016 | 111.9 | 1.489 | 0.8870 |
| **DBP** | 70.6 | 1.707 | 71.957 | 2.477 | 0.6506 |
| **PR** | 86.071 | 2.143 | 81.142 | 1.832 | 0.0826 |
| **RR** | 21.457 | 0.315 | 19.771 | 0.820 | 0.0541 |

The trend of C-reactive protein in the group receiving herbal supplements decreased from the first to the fourth day, but then increased from the fourth to the fifth day, and then decreased until the seventh day. But the control group in comparison to the intervention group had a significantly lower decrease in CRP in 7 days (P<0.05).

The mean number of hospitalization days in patients in the herbal supplement group was significantly lower than the patients in the control group. Patients in the herbal supplement group were hospitalized for 4.12 days and patients in the control group were hospitalized for 8.37 days (Table 4). ICU admission and death only happened in 3 (8.6%) patients of the control group.

**Table 4.** hospitalization time in intervention and control group

|  | Control<br>(n=35) | Intervention<br>(n=37) | P |
| --- | --- | --- | --- |
| Days of hospitalization, Mean ± SD | 8.37±2.71 | 4.12±1.55 | 0.001 |
| ICU admission, n(%) | 3 (8.6%) | 0 (0%) | 0.110 |
| Death, n(%) | 3 (8.6%) | 0 (0%) | 0.110 |

## Discussion

The results of this study showed that the addition of the proposed supplement (the combination of sugarcane, black myrobalan, and mastic) along with the treatment protocol of the Ministry of Health, can shorten the duration of treatment in patients with new coronavirus and relieve symptoms. While this study is the first study using this combination; no further studies with the same methodology and treatment method were available for comparison. Also, none of the herbs used in our combination was investigated in other studies as a treatment for COVID-19, except a clinical trial study in Iran, in which Anacyclus pyrethrum, Senna, Ferrula asafoetida, and Terminalia chebula effect have been planed to be tested on the COVID19 patients, but the study results are not yet reported. One of the main findings of this study was medication safety, as no subject showed any adverse events. This will help us to perform studies in a higher number of the patient or other groups of patients with more severe disease, underlying disease, and other age groups. The observed effects of the combination of sugarcane, black myrobalan, and mastic could be explained by the herb’s ingredients. In a 2008 study by Gupta et al. (2008), a randomized double-blind clinical trial of 60 febrile patients using aspirin (60 mg/kg body weight per day) as a standard drug compared with the use of sugarcane plant was done. this trial showed that fever after oral administration of sugarcane at a dose of 60 mg decreased rapidly and significantly, and this effect on fever was more stable and significant than aspirin. This may be the reason for the sooner fever control in our intervention group. But we were not able to monitor conventional antipyretic use in these patients and as a confounding factor, this was a limitation of our study; while for fever control, all patients had the same physician medical order.

The active ingredients of black myrobalan are terpenoids, carotenoids, flavonoids, alkaloids, tannins, and glycosides (Vemuri et al., 2019). In studies, this plant has been mentioned as a rich flavonoid plant (Sharma et al., 2019). While we did not have the opportunity of chemical constituent compounds analysis of herbs; previous administrations of black myrobalan were shown to be safe in humans. AyuFlex herbal, manufactured by Natreon Inc., New Jersey, USA, is a US FDA approved black myrobalan supplement (Murali et al., 2007). Black myrobalan has antiviral, antifungal, and antibacterial activity due to containing a variety of molecules. Black myrobalan has been reported to be an effective antiviral agent against swine flu type A, HSV-1, HIV-1 has been reported in various studies (Lopez et al., 2017; Ma et al., 2010). Other molecules in this plant are gallic acid and 3-glycol glucose molecules that inhibit the process of HIV-1 integration (Yukawa et al., 1996; Kim et al., 2001; Ardekani et al., 2011) and thus prevent viral infection without any side effects. Black myrobalan extract is effective in inhibiting the division of cytomegalovirus and is useful in people with immune deficiencies (Ahn et al., 2002; Nosalova et al., 2013; aik et al., 2004); The effect of using Black myrobalan has been shown against Respiratory syncytial virus, Hepatitis C virus, Herpes simplex virus, and Dengue virus (Jagtap et al., 1999). It also eliminates salivary bacteria by inhibiting the glycolysis pathway (Jagtap et al., 1999). Anti-inflammatory effects and improvement of asthma symptoms have been reported in the use of Terminalia chebula fruit extract. Animal studies showed that it can relieve cough even better than Codein, a confirmed medication for the cough (Nosalova et al., 2013; aik et al., 2004; Jagtap et al., 1999). Also, it’s anticaries properties may be as protection against tooth damage from daily sugarcane use, if the medication is going to be used for long periods (Sharma et al., 2011). But this hypothesis needs to be evaluated in further studies.

Pistacia lentiscus resina (mastics), the other component of our supplement, have antibacterial activity (Abidi et al., 2016), prevents inflammation due to its Linalool content (Shin et al., 2001). Studies show that mastics prevents the production of pro-inflammatory substances such as nitroxide and prostaglandin 2 (inhibition of cyclooxygenase 2 at mRNA and protein levels); Therefore, it is known as an anti-inflammatory and antioxidant substance (Peana et al., 2002; Mahmoudi et al., 2010). In laboratory studies on animal models, it has been shown that mastic plant can be effective in improving pulmonary fibrosis. the use of this plant extract in an animal model of asthma reduced airway inflammation by reducing the expression levels of TNF-α, IL-4, and IL-5, and improved pulmonary inflammation (Zhou et al., 2009). TNF-α is known to have potential activity in the cytokine storm, caused by COVID-19(Giamarellos-Bourboulis et al., 2020).

### Study limitations

Our study had some limitations. First due to a low number of subjects. Critically ill patients were not evaluated in this study. Also, patients with the underlying disease were excluded. Further researches could be conducted on these populations as no significant severe effect was recorded.

## Conclusion

The proposed combination of Sugarcane, Black Myrobalan, and Mastic seems to be effective in the symptom treatment and reducing the length of hospitalization in COVID-19 patients. Also, its safety was confirmed.

## Data Availability

All relevant and studied data are available in all versions of this manuscript.

## Conflict of interest

None.

## Abbreviations

ALT: Alanine aminotransferase
AHA: American Heart Association
AST: Aspartate aminotransferase
BUN: Blood urea nitrogen
BMI: Body mass index
Cr: Creatinine
DBP: Diastolic blood pressure
HB: Hemoglobin
INR: International normalized ratio
PTT: Partial thromboplastin time
PT: Prothrombin time
PR: Pulse rate
RBCS: Red blood cells
RR: Respiratory rate
SBP: Systolic blood pressure
TIM: Traditional Iranian Medicine
WBCS: White blood cells

## Figures and tables

**Flow diagram 1.**
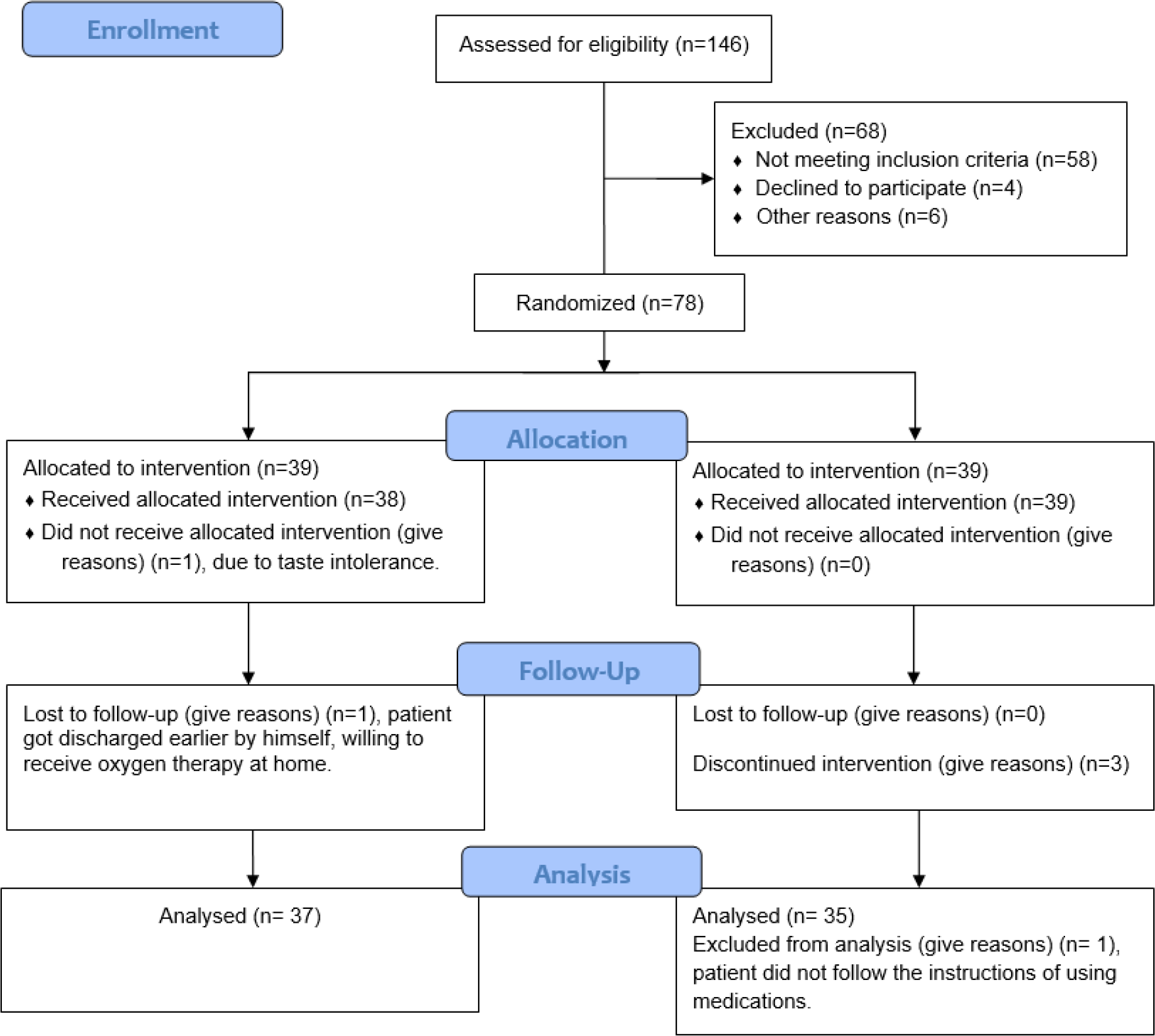
CONSORT flow chart

## Notes

### Competing Interest Statement

The authors have declared no competing interest.

### Clinical Trial

IRCT20200415047082N1

### Author Declarations

This study was approved by the ethics committee of Jahrom University of Medical Sciences under the ethical code IR.JUMS.REC.1399.003 and was registered in the Iranian registry of clinical trials under the number IRCT20200415047082N1.

## References

Abidi A, Beji RS, Kourda N, Ennigrou S, Ksouri R, Serairi RB, Jameleddine S, 2016. effect of Pistacia lentiscus oil on experimental pulmonary fibrosis effet de l’huile de Pistachier lentisque sur la fibrose pulmonaire expérimentale La Tunisie medicale 94(7).

Ahn M-J, Kim CY, Lee JS, Kim TG, Kim SH, Lee C-K, et al., 2002. Inhibition of HIV-1 integrase by galloyl glucoses from Terminalia chebula and flavonol glycoside gallates from Euphorbia pekinensis Planta Medica 68(05):457–9.

Aik G, Priyadarsini K, Naik D, Gangabhagirathi R, Mohan HJP,. 2004. Studies on the aqueous extract of Terminalia chebula as a potent antioxidant and a probable radioprotector Phytomedicine 11(6):530–8.

Ardekani MRS, Rahimi R, Javadi B, Abdi L, Khanavi MJJoTCM,. 2011. Relationship between temperaments of medicinal plants and their major chemical compounds J Tradit Chin Med31(1):27–31.

Belapurkar P, Goyal P, Tiwari-Barua P, 2014. Immunomodulatory effects of triphala and its individual constituents: a review Indian J of pharmaceu sci 76(6):467.

Cevik M, Bamford C, Ho A COVID-19 pandemic–A focused review for clinicians Clinical Microbiology and Infection Apr 25.

Dimas KS, Pantazis P, Ramanujam R, 2012. Chios mastic gum: a plant-produced resin exhibiting numerous diverse pharmaceutical and biomedical properties in vivo 26(5):777–85.

Giamarellos-Bourboulis EJ, Netea MG, Rovina N, Akinosoglou K, Antoniadou A, Antonakos N, Damoraki G, Gkavogianni T, Adami ME, Katsaounou P, Ntaganou M, Kyriakopoulou M, Dimopoulos G, Koutsodimitropoulos I, Velissaris D, Koufargyris P, Karageorgos A, Katrini K, Lekakis V, Lupse M, Kotsaki A, Renieris G, Theodoulou D, Panou V, Koukaki E, Koulouris N, Gogos C, Koutsoukou A, 2020. Complex Immune Dysregulation in COVID-19 Patients with Severe Respiratory Failure Cell Host Microb 27(6): 992–1000 e1003.

Gupta M, Shaw B, Mukherjee A, 2008. Evaluation of antipyretic effect of a traditional polyherbal preparation: A double-blind, randomized clinical trial Int J of pharmaco 4:190–5.

Jagtap AG, Karkera SG, 1999. Potential of the aqueous extract of Terminalia chebula as an anticaries agent J of Ethnopharmacology Dec 15;68(1-3):299–306.

Jin Y, Liang H, Cao C, Wang Z, Shu R, Li X, 1981. Immunological activity of bagasse polysaccharides (author’s transl) Zhongguo yao li xue bao= Acta pharmacologica Sinica 2(4):269.

Kim TG, Kang SY, Jung KK, Kang JH, Lee E, Han HM, et al & merge;Antiviral activities of extracts isolated from Terminalis chebula Retz, Sanguisorba officinalis L, 2001. Rubus coreanus Miq, and Rheum palmatum L against hepatitis B virus Phytotherapy res 15(8):718–20.

Ledon N, Casaco A, Rodriguez V, Cruz J, Gonzalez R, Tolon Z, et al, 2003. Anti-inflammatory and analgesic effects of a mixture of fatty acids isolated and purified from sugar cane wax oil Planta medica 69(04):367–9.

Li Y, Liu X, Guo L, Li J, Zhong D, Zhang Y, Clarke M, Jin R, 2020. Traditional Chinese herbal medicine for treating novel coronavirus (COVID-19) pneumonia: protocol for a systematic review and meta-analysis System rev Dec;9:1–6.

Lopez H, Habowski S, Sandrock J, Raub B, Kedia A, Bruno E, et al, 2017. Effects of dietary supplementation with a standardized aqueous extract of Terminalia chebula fruit (AyuFlex®) on joint mobility, comfort, and functional capacity in healthy overweight subjects: a randomized placebo-controlled clinical trial BMC comp and altern med 17(1):475.

Ma H, Diao Y, Zhao D, Li K, Kang T, 2010. A new alternative to treat swine influenza A virus infection: extracts from Terminalia chebula Retz Afr J Microbiol Res 4(6):497–9.

Maguire BJ, Guérin PJ, 2020. A living systematic review protocol for COVID-19 clinical trial registrations Wellcome Open Res. 5.

Mahmoudi M, Ebrahimzadeh MA, Nabavi SF, Hafezi S, Nabavi SM, Eslami Sh, 2010 Antiinflammatory and antioxidant activities of gum mastic Eur Rev Med Pharmacol Sci Sep; 14(9):765–9 PMID: 21061835.

Murali Y, Anand P, Tandon V, Singh R, Chandra R, Murthy P Long-term effects of Terminalia chebula Retz, 2007. on hyperglycemia and associated hyperlipidemia, tissue glycogen content and in vitro release of insulin in streptozotocin induced diabetic rats Experiment and clinic endocrinol & diabetes 115(10):641–6.

Nosalova G, Jurecek L, Chatterjee UR, Majee SK, Nosal S, Ray B, 2013. Antitussive activity of the water-extracted carbohydrate polymer from Terminalia chebula on citric acid-induced cough Evid-Base Comp and Alter Med Jan 1;2013.

Park M, Cook AR, Lim JT, Sun Y, Dickens BL., 2020. A systematic review of COVID-19 epidemiology based on current evidence J of Clinic Med 2020 Apr; 9(4):967.

Peana AT, D’Aquila PS, Panin F, Serra G, Pippia P, Moretti MD, 2002. Anti-inflammatory activity of linalool and linalyl acetate constituents of essential oils Phytomedicine Jan 1; 9(8):721–6.

Saleem A, Husheem M, Härkönen P, Pihlaja K, 2002. Inhibition of cancer cell growth by crude extract and the phenolics of Terminalia chebula retz fruit J of Ethnopharmacology. 81(3):327–36.

Sharma P, Prakash T, Kotresha D, Ansari MA, Sahrm UR, Kumar B, et al., 2011. Antiulcerogenic activity of Terminalia chebula fruit in experimentally induced ulcer in rats 49(3):262–8.

Sharma S, Singh B, Kumar H, 2019. A Critical Review of Pharmacological Actions of Haritaki (Terminalia chebula Retz) In Classical Texts J of Ayurveda and Integ Med Sci 4(4):258–69.

Shin T, Jeong H, Kim D, Kim S, Lee J, Chae B, et al., 2001. Inhibitory action of water soluble fraction of Terminalia chebula on systemic and local anaphylaxis J Ethnopharmacol 74(2):133–40.

Singh A, Lal UR, Mukhtar HM, Singh PS, Shah G, Dhawan RK, 2015. Phytochemical profile of sugarcane and its potential health aspects Pharmacognosy rev 9(17):45.

Singh G, Kumar P, 2013. Extraction, gas chromatography–mass spectrometry analysis and screening of fruits of Terminalia chebula Retz for its antimicrobial potential Pharmacognosy rese 5(3):162.

Vemuri PK, Dronavalli L, Nayakudugari P, Kunta A, Challagulla R, 2019. Phytochemical Analysis and Biochemical Characterization f Terminalia Chebula Extracts For its Medicinal use Biomed and Pharmaco J 12(3):1525–9.

Yukawa TA, Kurokawa M, Sato H, Yoshida Y, Kageyama S, Hasegawa T, et al., 1996. Prophylactic treatment of cytomegalovirus infection with traditional herbs Antiviral res 32(2):63–70.

Zhou L, Satoh K, Takahashi K, Watanabe S, Nakamura W, Maki J, Hatano H, Takekawa F, Shimada C, Sakagami H, 2009. Re-evaluation of anti-inflammatory activity of mastic using activated macrophages In Vivo Jul-Aug; 23(4):583–9 PMID: 19567394.

